# SARS-CoV-2-specific humoral and cellular immunity in renal transplant and haemodialysis patients treated with convalescent plasma

**DOI:** 10.1101/2020.12.09.20239673

**Authors:** Monika Lindemann, Adalbert Krawczyk, Sebastian Dolff, Margarethe Konik, Hana Rohn, Maximillian Platte, Laura Thümmler, Sina Schwarzkopf, Leonie Schipper, Maren Bormann, Lukas van de Sand, Marianne Breyer, Hannes Klump, Dietmar Knop, Veronika Lenz, Christian Temme, Ulf Dittmer, Peter A. Horn, Oliver Witzke

## Abstract

**Background:** When patients with chronic kidney disease are infected with severe acute respiratory syndrome coronavirus 2 (SARS-CoV-2) they can face two specific problems: Virus-specific immune responses may be impaired and remdesivir, an antiviral drug described to shorten the time to recovery, is contraindicated. Antiviral treatment with convalescent plasma could be an alternative treatment option.

**Methods:** In this case series we present two kidney transplant recipients and two patients dependent on haemodialysis who were infected with SARS-CoV-2 and received convalescent plasma. Antibodies against the spike 1 protein of SARS-CoV-2 were determined sequentially by IgG ELISA and neutralization assay and specific T cell responses by interferon-gamma ELISpot.

**Results:** Prior to treatment, in three patients antibodies were undetectable by ELISA (ratio < 1.1), corresponding to low neutralizing antibody titers (≤ 1:40). One patient was also negative to the ELISpot and two showed weak responses. After convalescent plasma treatment we observed an increase of SARS-CoV-2-specific antibodies (IgG ratio and neutralization titer) and of specific T cell responses. After intermittent clinical improvement one kidney transplant recipient again developed typical symptoms at day 12 after treatment and received a second cycle of convalescent plasma treatment. Altogether, three patients clinically improved and could be discharged from hospital. However, one multimorbid female in her early eighties deceased.

**Conclusions:** Our data suggest that the success of convalescent plasma therapy may only be temporary in patients with chronic kidney disease; which requires an adaptation of the treatment regimen. Close monitoring after treatment is needed for this patient group.

## Introduction

In patients with chronic kidney disease and infected with severe acute respiratory syndrome coronavirus 2 (SARS-CoV-2) treatment can be complicated because their immune function is suppressed due to medication to prevent allograft rejection and/or the underlying kidney disease. Thereby, the formation of specific antibodies and of T cell immunity is impaired; which can result in a prolonged persistence of SARS-CoV-2 (for up to two months^1^). Furthermore, remdesivir, an antiviral nucleoside analogue which shortened the time to recovery in adults hospitalized with coronavirus 2019 (COVID-19) disease^2^, is contraindicated in this special cohort. Antiviral treatment with convalescent plasma (CP) could be an alternative treatment option. Data on patients with chronic kidney disease infected with SARS-CoV-2 and receiving CP treatment are still limited. We are aware of only 12 described kidney transplant recipients who received CP.^3–6^ Whereas clinical improvement after CP has been shown for all six kidney transplant recipients included in three studies^3–5^, in the fourth study^6^ a mortality rate for solid organ recipients (including six with kidney allograft) in the range of recipients without CP treatment^7–9^ was reported (23%^6^ vs. 24-32%^7–9^, respectively). However, the previous reports did not present data on the course of SARS-CoV-2-specific antibodies or T cells in the patients.

It was the aim of the current study to follow-up up virus-specific humoral and cellular immunity in patients with chronic kidney disease who were infected with SARS-CoV-2 and received CP therapy. We functionally analysed the antibodies (by neutralisation assay) and measured T cell responses by the highly sensitive ELISpot method, using various protein antigens of SARS-CoV-2 as specific stimuli. Finally, in one transplant recipient who again developed typical COVID-19 symptoms after initial clinical improvement we had the chance to modify the treatment regimen and to apply a second cycle of CP therapy.

## Materials and Methods

### Patients and blood donors

The current case series includes two renal transplant recipients and two haemodialysis patients (Table 1) and their respective CP donors. One patient suffered from moderate and three from severe COVID-19 disease.^10^ All patients had chronic kidney disease according to the estimated glomerular filtration rate of 7-29 mL/min/1.73 m^2^. One cycle of CP consisted of three units, each containing 200-280 mL, which was applied at day 1, 3 and 5. The study was approved by the institutional review board of the University Hospital Essen, Germany (20-9256-BO for the patients and 20-9225-BO for the donors) and the study participants provided written informed consent. All necessary patient/participant consent has been obtained and the appropriate institutional forms have been archived. The procedures were in accordance with the institutional and national ethical standards as well as with the Helsinki Declaration of 1975, as revised in 2013.

**Table 1.** Clinical characteristics of patients with chronic kidney disease.

| **ID** | **Sex / Age<sup>a</sup> / Blood group** | **CP interval<sup>b</sup> (days)** | **CP units / cycles** | **Co-morbidity / cause of death** | **Severity of COVID-19 disease / Outcome** |
| --- | --- | --- | --- | --- | --- |
| RTX01 | F / 60's / O | 3 | 6 / 2 | 2 prior RTX, chronic antibody-mediated rejection, hypertension, asthma bronchiale | Severe<sup>c</sup> / A |
| RTX02 | F / 60's / A | 13 | 3 / 1 | RTX 2 weeks prior to SARS-CoV-2 infection, steroid-induced diabetes, hypertension | Moderate (CT: pneumonia, but clinically asymptomatic) / A |
| HD01 | F / 80's / A | 4 | 2 / 1 | HD since 2012, type II diabetes, coronary heart disease, apoplexy, acute event of fall / COVID-19 pneumonia | Severe<sup>c</sup> / D |
| HD02 | F / 70's / O | 7 | 3 / 1 | HD since 2020, type II diabetes, hypertension, chronic obstructive pulmonary disease, adipositas | Severe<sup>c</sup> / A |

<sup>a</sup>The precise age was replaced with an age range.
<sup>b</sup>Interval between onset of symptoms or positivity to SARS-CoV-2 PCR and treatment with convalescent plasma (CP)
<sup>c</sup>Oxygen supplementation but no mechanical ventilation
RTX, renal transplantation; HD, haemodialysis; CT, computer tomogram; A, alive D; dead.

## Methods

### Antibody ELISA

To assess SARS-CoV-2-specific humoral immunity, IgG antibodies were determined by a CE marked Anti-SARS-CoV-2 IgG semi-quantitative ELISA (Euroimmun, Lübeck, Germany), according to the manufacturer’s instructions. The ELISA plates were coated with recombinant SARS-CoV-2 spike (S) 1 protein (receptor binding domain). Serum samples were analysed automatically at a 1:100 dilution, using the Immunomat^™^ (Virion\Serion, Würzburg, Germany). Results are given as ratio (patient sample / control sample). An antibody ratio of ≥ 1.1 was considered positive, of ≥ 0.8 to < 1.1 borderline and of < 0.8 negative.

### Virus neutralization assay

The function of specific antibodies was measured by a cell-culture based neutralization assay, using Vero E6 cells (ATCC® CRL-1586™, Manassas, Virginia, USA) and a clinical isolate of SARS-CoV-2.^11,12^ Neutralization capacity was determined by endpoint dilution assay, expressed as 50% tissue culture infective dose (TCID_50_)/mL. Serial dilutions (1:20 to 1:1280) of the respective sera were pre-incubated with 100 TCID_50_ of SARS-CoV-2 for one hour at 37°C and added afterwards to confluent Vero E6 cells cultured in 96-well microtiter plates. On day 3 after infection, the cells were stained with crystal violet (Roth, Karlsruhe, Germany) solved in 20 % methanol (Merck, Darmstadt, Germany) and the appearance of cytopathic effects (CPE) was analysed by light microscopy. The neutralizing titer was defined as the reciprocal of the highest serum dilution at which no CPE breakthrough in any of the triplicate cultures was observed.

### ELISpot assay

To assess SARS-CoV-2-specific cellular immunity, we performed ELISpot assays, using peptide pools of the S1/S2 protein, of the S1 protein and of the membrane (M) protein (Miltenyi Biotec, Bergisch Gladbach, Germany) and an S1 protein antigen of SARS-CoV-2 (Sino Biological, Wayne, PA, U.S.A.). We tested 250,000 peripheral blood mononuclear cells (PBMC) per cell culture and measured IFN-γ production after 19 hours, as published recently in detail^12^ (https://wwwnc.cdc.gov/eid/article/27/1/20-3772-app1.pdf). Spot numbers were analysed by an ELISpot reader (AID Fluorospot, Autoimmun Diagnostika GmbH, Strassberg, Germany). Mean values of duplicate cell cultures were considered. SARS-CoV-2-specific spots were determined as stimulated minus non-stimulated (background) values (spots increment). We defined threefold higher SARS-CoV-2-specific spots versus background together with at least three spots above background as positive response. This cut-off was set based on negative control values as described previously.^12^

## Results

In three patients with undetectable SARS-CoV-2-specific IgG (ratio <1.1) and low neutralizing antibody titers (≤ 1:40) we observed an increase of antibody titers (Figure 1A-C). One patient who was transplanted twice (RTX01) initially showed a clinical response to CP therapy, but at day 12 again developed typical symptoms of COVID-19 disease (fever and shortage of air). Therefore, she received another cycle of CP therapy (from the same donor). SARS-CoV-2 antibodies increased after both CP cycles and SARS-CoV-2 viral load decreased (ct value to the PCR increased from 17.8 to 25.8 after the first and to 34.9 after the second CP cycle). The antibody ratios in the first three patients prior to the CP therapy were 0.15, 0.14 and 0.17 and the respective neutralizing antibody titers 1:20, < 1:20 and 1:40. After CP therapy, antibodies in the patients reached a maximum ratio of 3.07, 2.19 and 3.70, corresponding to a neutralizing titer of up to 1:640, 1:160 and 1:640, respectively. In the donors, the antibody ratios were 5.83, 7.33 and 10.44 and the neutralizing titers 1:1280, 1:320 and 1:160, respectively. The fourth patient with pre-existing antibodies showed an increase of specific immunity (ratio 5.96 → 7.01; neutralizing titer 1:640 → 1:1280) (Figure 1D). SARS-CoV-2-specific antibodies in the CP donor of the fourth patient were lower (ratio 3.39, neutralizing titer 1:320). Cellular immunity could be followed-up by IFN-γ ELISpot in three patients, using four different SARS-CoV-2-specific antigens (peptide pools of the S1/S2, S1 and M protein and an S1 protein antigen). In all three patients IFN-γ production to the ELISpot intermittently increased after CP therapy, reaching a maximum at day 6 to 14 after therapy. These three patients clinically improved and could be discharged from hospital. However, one multimorbid female in her early eighties (HD01) deceased due to COVID-19 pneumonia on day 4 after initiation of CP therapy.

**Figure 1.**
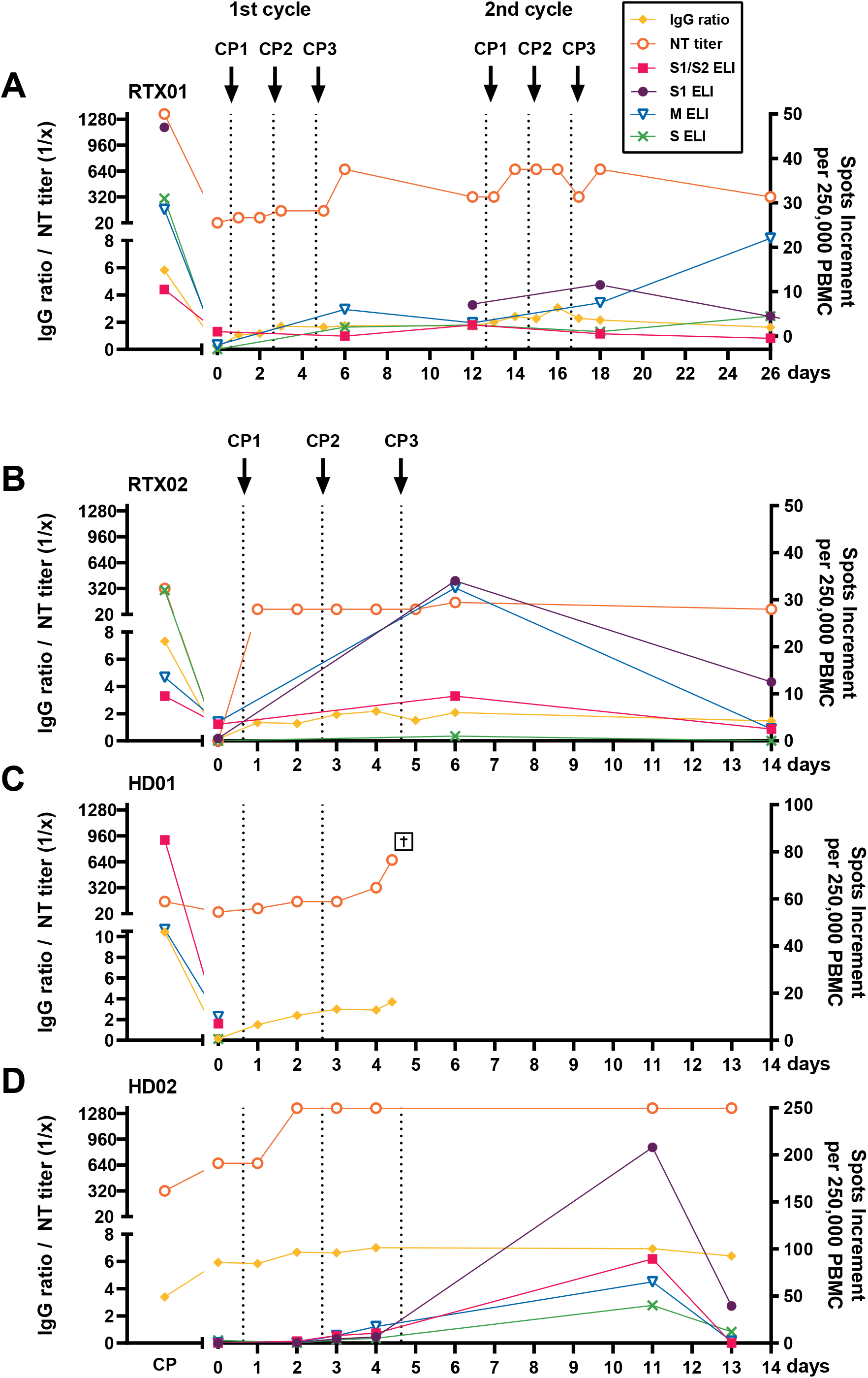
Course of specific humoral and cellular immunity in four patients with chronic kidney disease infected with SARS-CoV-2 and receiving convalescent plasma treatment. Antibodies were determined by an S1 specific IgG ELISA (Euroimmun) and by cell-culture based neutralization assay (NT titer). T cell responses were analysed by an interferon-gamma (IFN-γ) ELISpot assay, using peptide pools of the S1/S2, S1 and M protein and an S1 protein antigen as specific stimuli (depicted as S1/S2, S1, M and S ELI). We here present data on two kidney transplant recipients (RTX01, RTX02) and two patients on haemodialysis (HD01, HD02) and compared their immune responses with those of the corresponding donors of convalescent plasma (CP, shaded area). SARS-CoV-2-specific antibody data (IgG ratio and NT titer) are given on the left Y axis and ELISpot data on the right one. Vertical dotted lines indicate the time points of convalescent plasma applications (CP1, CP2 and CP3). Related data points are connected.

## Discussion

Our data indicate that CP therapy led to an increase of specific humoral and cellular immunity in two kidney transplant recipients and two haemodialysis patients with SARS-CoV-2 infection. CP therapy could bridge the phase of acute COVID-19 disease. However, presumably due to drug-induced immunosuppression or impaired kidney function, the immune responses were not as long-acting as expected. In one patient with two prior kidney transplantations (RTX01) two cycles of therapy were necessary for successful treatment. It can be supposed that the patient herself was unable to mount an adequate antibody response and that the passively transferred antibodies partly bound the virus that resides in the affected organs and in the respective lymphoid tissue.^13^ Theoretically, it is possible that CP therapy mitigates the native humoral immune response and leaves an individual vulnerable to subsequent reinfection with SARS-CoV-2.^3,14^ This phenomenon appears more likely in immunosuppressed vs. otherwise healthy individuals. Concerning ELISpot data, we observed a maximum of IFN-γ responses shortly after completion of the CP cycle. Of note, cellular immunity is regarded as important for recovery from SARS-CoV-2 infection^15^ and appears as short-lived in the current cohort. As CP therapy is a form of passive immunization, an increase of cellular responses is not expected at first glance. It can be speculated that due to the reduction of the viral load the patients were able to generate specific T cell responses by themselves. It was previously described that high viral load (of the hepatitis B virus) caused antigen-specific immune tolerance.^16^ Possibly, a similar imbalance between viral load and control by the immune system occurred in the current cohort. After an initial increase IFN-γ production decreased again, which could reflect the fact that due to the decrease of antibodies antigen-specific immune tolerance re-appeared or that pro-inflammatory immune responses shifted to anti-inflammatory responses.^17^ Moreover, treatment with tacrolimus, mycophenolate mofetil and prednisone (in both transplant recipients) and toxic metabolites due to chronic kidney disease suppressed T cell function, which could impede long-term protection against reinfection.^3,18^

Both kidney transplant recipients showed clinical improvement and three out of four patients with chronic kidney disease; which is in the range of previous reports^3–6^. However, due to the low patient number, it was beyond the aim of our study to answer the question if CP therapy was effective. This answer can only be given by large randomized clinical studies such as the Randomised Evaluation of COVID-19 Therapy (RECOVERY) trial ^19^; which are currently underway.

In conclusion, our data suggest that despite an increase of SARS-CoV-2-specific immunity the success of CP therapy may only be temporary in a subset of patients with chronic kidney disease. Thus, short-term treatment control appears mandatory for this patient group. If necessary, the the treatment regimen has to be adapted.

## Data Availability

Data are available on request from the corresponding author.

## Acknowledgements

The authors would like to thank Babette Große-Rhode and Martina Filipovic for their excellent technical assistance. We furthermore thank all volunteers for their participation and the donation of blood samples.

## Abbreviations

SARS-CoV-2: severe acute respiratory syndrome coronavirus 2
CP: convalescent plasma
COVID-19: coronavirus 2019
IFN-γ: interferon-gamma
PBMC: peripheral blood mononuclear cells
S1: spike 1

## Notes

**Disclosure**: The authors declare no conflicts of interest.

**Funding**: This work was supported by the Stiftung Universitätsmedizin Essen (A.K.) and the Rudolf Ackermann Foundation (O.W.).

### Competing Interest Statement

The authors have declared no competing interest.

### Funding Statement

This work was supported by the Stiftung Universitaetsmedizin Essen (A.K.) and the Rudolf Ackermann Foundation (O.W.).

### Author Declarations

The study was approved by the institutional review board of the University Hospital Essen, Germany (20-9256-BO for the patients and 20-9225-BO for the donors) and the study participants provided written informed consent. All necessary patient/participant consent has been obtained and the appropriate institutional forms have been archived.

